# Magnitude and kinetics of anti-SARS-CoV-2 antibody responses and their relationship to disease severity

**DOI:** 10.1101/2020.06.03.20121525

**Authors:** Kara L Lynch, Jeffrey D Whitman, Noreen P Lacanienta, Erica W Beckerdite, Shannon A Kastner, Brian R Shy, Gregory M Goldgof, Andrew G Levine, Sagar P Bapat, Susan L Stramer, Jonathan H Esensten, Allen W Hightower, Caryn Bern, Alan HB Wu

**Author notes:** Correspondence to: Kara L. Lynch, PhD, University of California, Department of Laboratory Medicine, 1001 Potrero Ave. Bldg. 5 2M16, San Francisco, CA 94110.

## Abstract

**Background:** SARS-CoV-2 infection can be detected indirectly by measuring the host immune response. Anti-viral antibody concentrations generally correlate with host protection and viral neutralization, but in rare cases, antibodies can promote disease progression. Elucidation of the kinetics and magnitude of the SARS-CoV-2 antibody response is essential to understand the pathogenesis of COVID-19 and identify potential therapeutic targets.

**Methods:** Sera (n=533) from patients with RT-PCR confirmed COVID-19 (n=153) were tested using a high-throughput quantitative IgM and IgG assay that detects antibodies to the spike protein receptor binding domain and nucleocapsid protein. Individual and serial samples covered the time of initial diagnosis, during the disease course, and following recovery. We evaluated antibody kinetics and correlation between magnitude of the response and disease severity.

**Results:** Patterns of SARS-CoV-2 antibody production varied considerably. Among 52 patients with 3 or more serial specimens, 44 (84.6%) and 42 (80.8%) had observed IgM and IgG seroconversion at a median of 8 and 10 days, respectively. Compared to those with milder disease, peak measurements were significantly higher for patients admitted to the intensive care unit for all time intervals between 6 and 20 days for IgM, and all intervals after 5 days for IgG.

**Conclusions:** High sensitivity assays with a robust dynamic range provide a comprehensive picture of host antibody response to SARS-CoV-2. IgM and IgG responses were significantly higher in patients with severe than mild disease. These differences may affect strategies for seroprevalence studies, therapeutics and vaccine development.

## Introduction

Serological testing is widely proposed as a major tool to manage the COVID-19 pandemic, playing a key role in more accurate disease burden assessment, identification of potential donors for therapeutic immune plasma, and tracking evolution toward population level immunity.^1,2^ Nearly all patients with symptomatic infection develop detectable IgM and IgG antibodies within several weeks of symptom onset, consistent with patterns seen in other systemic viral infections.^3-5^ Although detectable IgM usually precedes IgG, some patients show simultaneous rises in both antibodies, and the intensity of responses is heterogeneous.^6^ However, most published COVID-19 data derive from hospitalized patients.^3,5,6^ Sparse evidence suggests that mild or asymptomatic infection may result in substantially lower IgG responses.^6,7^ If confirmed, this feature carries major implications for population assessments and identification of convalescent plasma donors. In the small number of patients studied to date, the magnitude of the IgG response correlated with that of neutralizing antibodies, suggesting that not all serological responses are equivalent in terms of future protection or the ability to transfer protective antibodies.^3,8,9^ Accurate quantitative understanding of anti-SARS-CoV2 antibody responses will be essential both for public health interventions and therapeutic applications.^2,10^

Using a clinically-validated high-throughput assay that provides quantitative IgM and IgG results, we report the evolution of antibody responses, and compare the magnitude of convalescent antibody responses to patients with critical and non-critical COVID-19 disease.

## Methods

### Ethical review

Two separate protocols, one for Zuckerberg San Francisco General Hospital (ZSFG) remnant specimens (IRB #20-30387) and the other for convalescent plasma donor screening (IRB #20-30637), were approved by the Institutional Review Board of the University of California, San Francisco. The committee judged that written consent was not required for use of remnant specimens. Written informed consent was obtained for convalescent plasma donor screening.

### Subjects and specimens

The primary cohort analysis utilized remnant serum or plasma samples from routine clinical laboratory testing at ZSFG hospital. All patients had positive results by SARS-CoV-2 real-time polymerase chain reaction (RT-PCR) in nasopharyngeal swabs. Clinical data were extracted from electronic health records and included demographic information, major co-morbidities, patient-reported symptom onset date, symptoms and indicators of disease severity. Analyses of time intervals 3 weeks or more after symptom onset included an additional set of serum samples from patients who were screened for convalescent plasma donation. COVID-19 convalescent plasma donors were recruited via medical record searches and public appeals. Potential donors over 18 years of age with a self-reported positive SARS-CoV-2 RT-PCR test result were screened for allogeneic blood donation eligibility. A serum sample was collected by phlebotomy for SARS-CoV-2 antibody testing.

### Antibody measurement

Antibodies to SARS-CoV-2 (IgM and IgG) were measured using the Pylon 3D automated immunoassay system (ET Healthcare, Palo Alto, CA) as described previously.^11^ In brief, quartz glass probes pre-coated with either affinity purified goat anti Human IgM (IgM capture) or Protein G (IgG capture) are dipped into diluted patient sample (15 uL). Following a wash sequence, the probe is dipped into the assay reagent containing both biotinylated recombinant spike protein receptor binding domain (RBD) and nucleocapsid protein (NP). After a wash sequence, the probe is incubated with a Cy®5-streptavidin (Cy5-SA) polysaccharide conjugate reagent. The polysaccharide carries multiple copies of Cy5-SA allowing for cyclic amplification of the fluorescence signal. The background corrected signal was reported as relative fluorescent units (RFU) which is proportional to the amount of specific antibodies in the sample.

### Antibody measurement method validation

Eighty American Red Cross blood donor samples collected prior to June 2018 were analyzed, and the mean and standard deviation calculated for the distribution of IgM and IgG readings. Cut-offs that represented the mean plus multiples of the standard deviation were applied to 71 samples collected from febrile patients with upper respiratory symptoms that were either SARS-CoV-2 RT-PCR negative or positive for other respiratory viruses (BioFire® FilmArray® Respiratory 2 Panel, Salt Lake City, UT) including, coronaviruses (HKU1, 229E, OC43), influenza virus A (H1, H3, HI 2009), human rhinovirus/enterovirus, human metapneumovirus, respiratory syncytial virus, parainfluenza virus 1 and 4, and adenovirus. The mean plus 4 standard deviations provided 98.6% and 100% specificity for IgM and IgG respectively and was used as the cut-off for all subsequent analyses. Intra- and inter-day imprecision (5 samples per level per day for 5 days) for quality control patient pools (negative, low positive and high positive) was <20% for both IgM and IgG. Our validations studies confirmed that results were not affected by interferences such as lipemia, bilirubinemia, and hemolysis. A SARS-CoV-2 human IgG standard spiked into negative human serum was measured at 6 concentrations ranging from 1 to 300 μg/mL and provided a linear response with 300 μg/mL corresponding to 6976 RFU. High patient samples (n=3) for IgM and IgG were serially diluted allowing for verification of the analytical measurement range from the cutoff to 1900 RFU for IgM and the cutoff to 7000 RFU for IgG.

### Statistical analysis

We categorized patients based on their level of care; patients admitted to an intensive care unit at any time were classified as ICU patients, whereas those admitted to a hospital ward or managed as outpatients were considered non-ICU patients. Differences in categorical variables were evaluated by Chi Square test. Continuous variables were analyzed using the Wilcoxon Rank Sum or Kruskal-Wallis test with t-approximation or exact methods, as appropriate. For analyses within the primary cohort, we computed maximum antibody responses for 5-day time intervals, using the peak measurement during the time interval for each person. We estimated the median time in days from symptom onset to seroconversion using Kaplan-Meier survival analyses. For the analysis that included specimens collected under the convalescent plasma donor protocol, we compared antibody responses at 21 days or more after symptom onset for ICU versus non-ICU patients. This analysis utilized the results of the last specimen collected from primary cohort patients to avoid a potential bias due to the cohort group having more readings and higher likelihood of a detected maximum than the non-severe convalescent group. All statistical tests were two-tailed. Analyses were performed in SAS version 9.4 (SAS Institute Inc., Cary, NC).

## Results

The primary cohort included 94 SARS-CoV2 RT-PCR-positive patients, 62 (66%) admitted to the hospital and 32 outpatients (Table 1). Of the hospitalized patients, 26 (41.9%) were admitted to the intensive care unit and 19 (30.6%) required mechanical ventilation. The primary cohort thus comprised 26 ICU patients and 68 non-ICU patients. Two-thirds were male and the median age was 49 years; ICU patients were slightly older than less severely ill patients. Seventy-one (77%) patients identified as Hispanic, consistent with the hospital’s patient population and emerging evidence on San Francisco’s COVID-19 demographics. Hypertension, type II diabetes and obesity were more common among ICU than non-ICU patients, but the difference was significant only for obesity. Reported symptoms were typical of COVID-19; only dyspnea was significantly more frequent among ICU patients than less severely ill patients.

**Table 1.** Demographic and clinical characteristics of the 94 primary cohort patients with covid-19 confirmed by SARS-CoV2 reverse transcriptase polymerase chain reaction (RT PCR).

| Characteristic<sup>1</sup> | All patients<br>N=94 | ICU patients<br>N=26 | Non-ICU patients<br>N=68 | P value<sup>2</sup> |
| --- | --- | --- | --- | --- |
| Age |  |  |  |  |
| Median [IQR] | 49 [39 – 58] | 53.5 [41 - 62) | 47.5 [38.5 – 57] | 0.13 |
| Mean (SD) | 49.4 (14.4) | 53.4 (14.1) | 47.9 (14.3) |  |
| Male sex | 64 (68.1) | 18 (69.2) | 46 (67.7) | 1.0 |
| Admitted to hospital | 62 (66.0) | 26 (100) | 36 (52.9) | <0.0001 |
| Mechanical ventilation | 19 (20.2) | 19 (73.1) | 0 (0) | <0.0001 |
| Medical history |  |  |  |  |
| Hypertension | 36 (38.3) | 14 (53.9) | 22 (32.4) | 0.10 |
| Type II diabetes | 37 (39.4) | 14 (53.9) | 23 (33.8) | 0.10 |
| Obesity | 19 (20.2) | 9 (34.6) | 10 (14.7) | 0.045 |
| Chronic renal disease | 6 (6.4) | 3 (11.5) | 3 (4.4) | 0.34 |
| HIV | 3 (3.2) | 0 (0) | 3 (4.4) | 0.56 |
| Organ transplantation | 2 (2.3) | 0 (0) | 2 (3) | 1.0 |
| Symptoms<sup>3</sup> |  |  |  |  |
| Fever | 76 (81.7) | 22 (84.6) | 54 (80.6) | 0.77 |
| Cough | 80 (86.0) | 22 (84.6) | 58 (86.6) | 0.75 |
| Sore throat | 23 (24.7) | 7 (26.9) | 16 (23.9) | 0.79 |
| Dyspnea | 62 (66.7) | 22 (84.6) | 40 (59.7) | 0.03 |
| Chest pain | 21 (22.6) | 7 (26.9) | 14 (20.9) | 0.58 |
| Disordered smell/taste | 4 (4.3) | 0 (0) | 4 (6.0) | 0.57 |
| Diarrhea | 13 (14.0) | 4 (15.4) | 9 (13.4) | 0.75 |
| Nausea or vomiting | 13 (14.0) | 1 (3.9) | 12 (17.9) | 0.10 |
| Headache | 29 (31.2) | 6 (23.1) | 23 (34.3) | 0.33 |
| Myalgia | 34 (36.6) | 9 (34.6) | 25 (37.3) | 1.0 |

<sup>1</sup>Medians [interquartile range], mean (standard deviation) or N (%) are reported.
<sup>2</sup>P value for comparison of ICU versus non-ICU patients by Fisher exact or Wilcoxon rank sum test
<sup>3</sup>Symptom data missing for one non-ICU patient

The primary cohort analysis included 474 remnant plasma or serum specimens. ICU patients contributed more specimens and had longer median follow-up time than non-ICU patients (Table 2). The rapidity of antibody rise and the shape of the curves varied considerably (Figure 1). Among all 94 patients, 56 (59.6%) and 46 (48.9%) developed detectable IgM and IgG antibodies, with median times to IgM and IgG seroconversion of 10 and 12 days, respectively. Follow-up times were 15 days or less for 33 (86.8%) and 42 (87.5%) of those without IgM or IgG seroconversion, respectively. Six individuals with specimens at least 16 days after symptom onset remained IgG seronegative. Of these, two had iatrogenic immunosuppression, one on mycophenolate, prednisone and tacrolimus post-renal transplant, and the other on adalimumab, prednisolone and cyclopentolate for reactive arthritis. The second patient showed IgG seroconversion after suspension of adalimumab for 40 days.

**Table 2.** Anti-SARS-CoV2 IgM and IgG results in specimens from the 94 covid-19 patients in the primary cohort.

|  | IgM |  |  | IgG |  |  |
| --- | --- | --- | --- | --- | --- | --- |
|  | All<br>N=94 | Non-ICU<br>N=68 | ICU<br>N=26 | All<br>N=94 | Non-ICU<br>N=68 | ICU<br>N=26 |
| All patients<sup>1</sup> |  |  |  |  |  |  |
| Median specimen number [IQR] | 3 [1 – 8] | 2 [1 – 3.5] | 11 [8 – 15] | 3 [1 – 8] | 2 [1 – 3.5] | 11 [8 – 15] |
| Last follow-up day | 11.5 [5 – 17] | 7 [3 – 13] | 25 [17 – 30] | 11.5 [5 – 17] | 7 [3 – 13] | 25 [17 – 30] |
| Seroconverted N (%) | 56 (59.6) | 33 (48.5) | 23 (88.5) | 46 (48.9) | 21 (30.9) | 25 (96.2) |
| Days to seroconversion, median<sup>2</sup> | 10 | 10 | 8 | 12 | 13 | 9 |
| 95% confidence interval | 8 – 12 | 9 – 13 | 7 – 13 | 9 – 14 | 10 – 25 | 8 – 14 |
| Interquartile range | 7 – 14 | 7 – 16 | 7 – 14 | 8 – 17 | 9 – 25 | 7 – 15 |
| Peak reading | 147.5 [46 – 682] | 85 [33 – 347] | 969 [457-1289] | 27.5 [7 – 817] | 10.5 [6 – 134] | 2577 [657 – 4096] |
| Days to peak | 10 [5 – 15] | 7 [3 – 13] | 16 [12 – 19] | 11.5 [5 – 17] | 7 [3 – 13] | 19.5 [13 – 28] |
| Patients with 3+ specimens | All<br>N=52 | Non-ICU<br>N=26 | ICU<br>N=26 | All<br>N=52 | Non-ICU<br>N=26 | ICU<br>N=26 |
| Median specimen number [IQR] | 7 [4 – 11] | 4 [3 – 5] | 11 [8 – 15] | 7 [4 – 11] | 4 [3 – 5] | 11 [8 – 15] |
| Last follow-up day | 15.5 [10.5 – 25.5] | 12 [8 – 15] | 25 [17 – 30] | 15.5 [10.5 – 25.5] | 12 [8 – 15] | 25 [17 – 30] |
| Seroconverted N (%) | 44 (84.6) | 21 (80.8) | 23 (88.5) | 42 (80.8) | 17 (65.4) | 25 (96.2) |
| Days to seroconversion, median<sup>2</sup> | 9 | 10 | 8 | 10 | 10 | 9 |
| 95% confidence intervals | 7 – 10 | 7 – 11 | 7 – 13 | 8 – 13 | 7 – 13 | 8 – 14 |
| Interquartile range | 6 – 13 | 6 – 12 | 7 – 14 | 7 – 15 | 7 – 25 | 7 – 15 |
| Peak reading | 582.5 [173 -1056] | 387 [129 – 620] | 969 [457 – 1289] | 642 [101 – 2576] | 259 [17 – 620] | 2577 [657 – 4096] |
| Days to peak | 13 [9 – 16.5] | 10.5 [7 – 13] | 16 [12 – 19] | 13.5 [10.5 – 25.5] | 11.5 [7 – 14] | 19.5 [13 – 28] |

<sup>1</sup>Medians [interquartile range] or N (%) are reported.
<sup>2</sup>Based on Kaplan-Meier analysis

**Figure 1.**
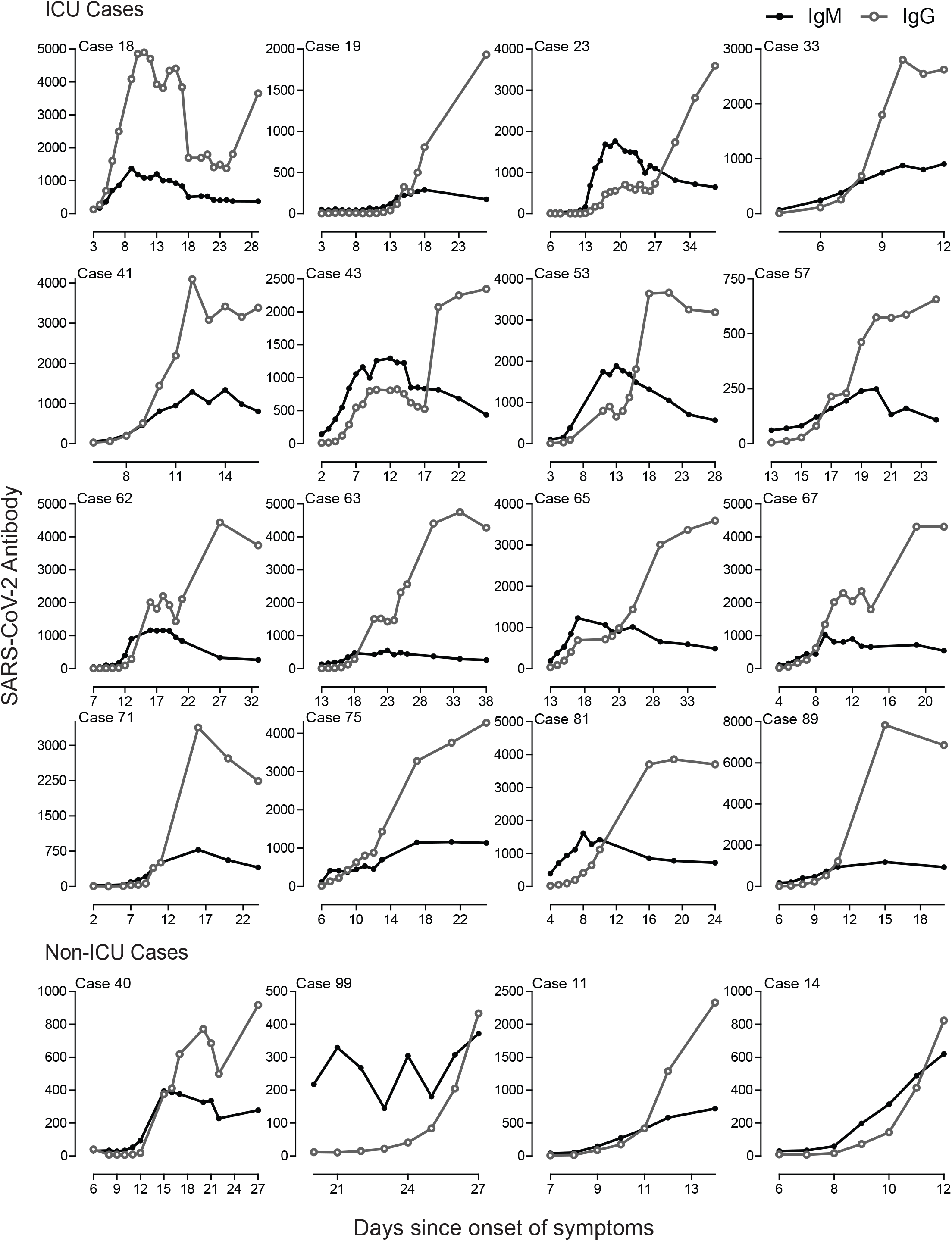
Kinetics of IgM and IgG responses for 20 hospitalized patients by days after symptom onset.

We restricted further Kaplan-Meier analyses to the 52 patients with 3 or more specimens, 26 non-ICU (24 inpatient and 2 ambulatory) and 26 ICU patients. In these analyses, 44 (84.6%) and 42 (80.8%) patients had observed IgM and IgG seroconversion, with median times to first positive specimen of 8 and 10 days, respectively. Peak IgM and IgG readings were significantly higher in specimens from ICU than non-ICU patients, but these analyses were confounded by the strong association between availability of later specimens and ICU status. To mitigate the effects of confounding, we compared peak IgM and IgG readings within 5-day time intervals. Peak readings were significantly higher for specimens from ICU than non-ICU patients for all time intervals between 6 and 20 days for IgM, and all intervals after 5 days for IgG (Figure 2A and 2B).

**Figure 2.**
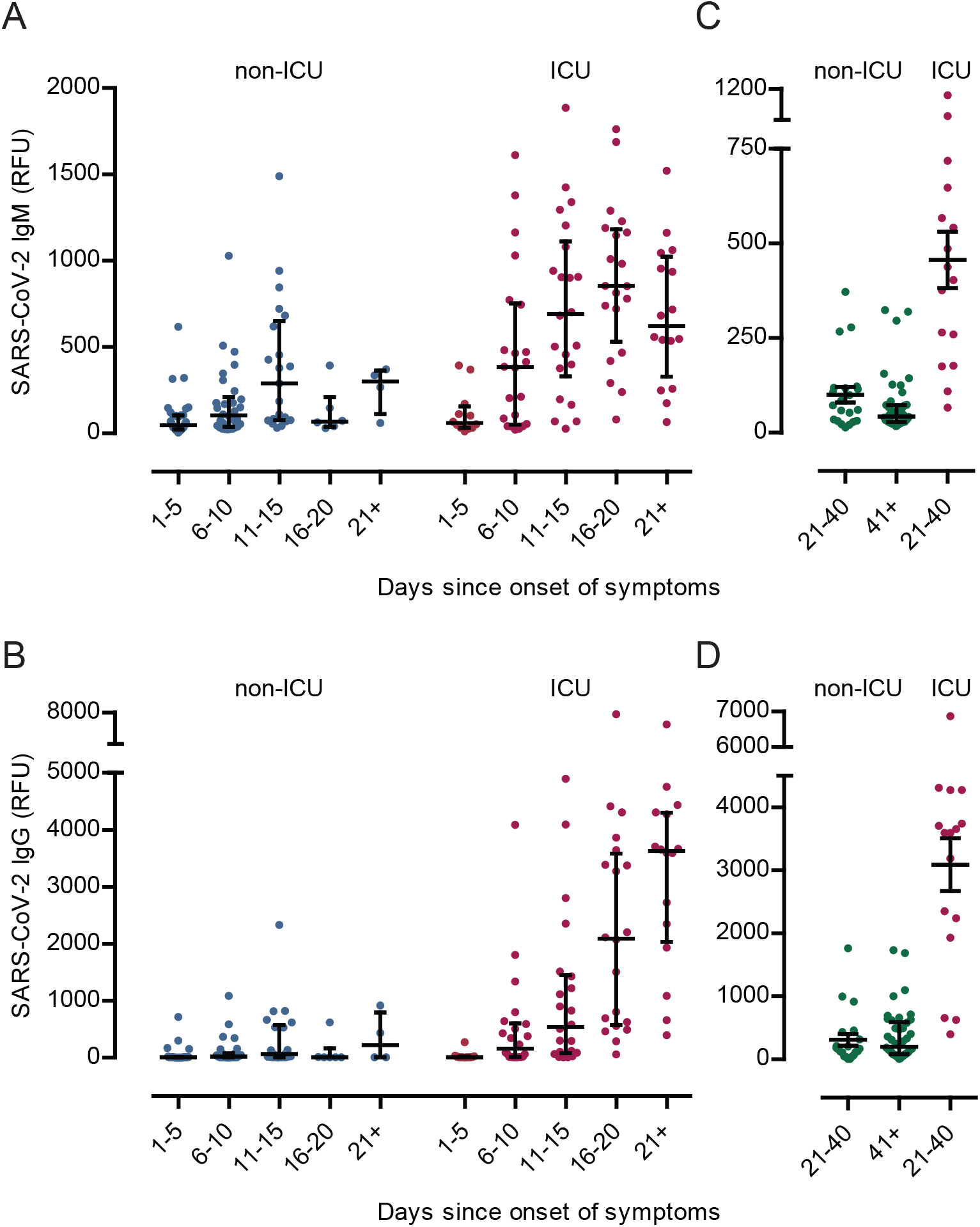
Distribution of IgM and IgG responses in relative fluorescence units (RFU) for specimens by level of care (ICU vs non-ICU). Panels A and B show results from hospital patients only (P values 0.34, 0.03, 0.03, 0.002 and 0.06 (IgM) and 0.05, 0.02, 0.02, 0.003, 0.01 (IgG) for 1-5, 6-10, 11-15, 16-20 and >21 days after symptom onset, respectively). Panels C and D include data from convalescent serum screening; includes 16 ICU patients, N=21 non-ICU patients with specimens at 21-40 days and 42 non-ICU patients with specimens at 41-70 days post onset (P values shown in Table 3).

We performed a series of additional analyses for samples collected at least 3 weeks after symptom onset (Table 3; Figures 2C and 2D). The specimens included in this analysis comprised the last collected specimens from 20 primary cohort patients, 16 ICU and 4 non-ICU, plus 59 specimens collected under the convalescent plasma donor protocol, all non-ICU. Convalescent sera from non-ICU patients were collected a median of 45 (range 21 – 70) days after symptom onset, significantly later than for ICU patients. We therefore performed stratified analyses for ICU patients compared to non-ICU patients with specimens collected 21-40 and 41-70 days after specimen onset. Within the 21-40 day interval, the timing of specimens for ICU and non-ICU samples was comparable. In all analyses, IgM and IgG concentrations were significantly higher for ICU than non-ICU patients. ICU patients were significantly older than non-ICU patients; in comparisons limited to patients 40 years or older, IgM and IgG responses remained significantly higher among ICU than non-ICU patients for all comparisons (data not shown).

**Table 3.** IgM and IgG responses among COVID-19 patients with specimens at 21 or more days after symptom onset. Includes 20 patients from the primary cohort and 63 patients recruited under the convalescent plasma donor protocol.

|  | ICU<br>(N=16) | Non-ICU, 21-40 days<br>(N=21)<sup>1</sup> | P value<br>(vs ICU) | Non-ICU, 41-70 days<br>(N=42)<sup>2,3</sup> | P value<br>(vs ICU) |
| --- | --- | --- | --- | --- | --- |
| Median age (years) | 57.5 [45 – 63.5] | 44 [27 – 58] | 0.03 | 38 [33 – 50] | 0.002 |
| Median [IQR] days after onset* | 28.5 [25 – 34] | 33 [28-38] | 0.15 | 48 [44 – 52] | <0.0001 |
| IgM positive, N (%)** | 15 (93.8) | 13 (61.9) | 0.05 | 18 (42.9) | 0.0004 |
| Median [IQR] IgM titer* | 421 [219– 607] | 72 [32 – 118] | 0.0001 | 43 [28 – 73] | <0.0001 |
| IgG positive, N (%)** | 16 (100) | 16 (76.2) | 0.06 | 38 (90.5) | 0.57 |
| Median [IQR] IgG titer* | 3595 [2086 – 4009] | 159 [71 – 317] | <0.0001 | 202 [84 – 579] | <0.0001 |

\*P-values computed using: Fisher Exact or Chi-Square test.
\*\*P-values computed using: Wilcoxon Rank Sum test.
<sup>1</sup>Non-ICU patients with specimens 21-40 days after symptom onset. Includes 17 treated as outpatients and 4 admitted to ward; four (3 inpatients, 1 outpatient) from the primary cohort, the others from convalescent plasma study
<sup>2</sup>Patients with specimens 41-70 days after onset; 41 outpatients and one admitted to ward, all from convalescent plasma study
<sup>3</sup>P>0.05 for comparison of non-ICU 21-40 days vs non-ICU 41-70 days for all variables except days since onset

## Discussion

The urgency to disseminate SARS-CoV-2 research data has resulted in papers with conflicting data regarding the host humoral response to the virus. Reported median time to seroconversion has ranged from as early as 4 days to as late as 14 days.^5,6,12,13^ A significant association between disease severity and antibody responses was described in some publications^5,6^ but not others.^3^ The incomplete nature of the data available to date has impeded a full elucidation of antibody development after infection and robust statistical analyses of differences between clinical groups. In this study, the serial nature of the samples evaluated from the time of RT-PCR testing to hospital discharge allowed for the direct observation of seroconversion in 42 patients for IgM and 44 for IgG. Median times to IgM and IgG seroconversion were 8 and 10 days, respectively. The rate and magnitude of the antibody response varied between individuals, but peak IgM and IgG levels were significantly associated with disease severity in nearly all time intervals in the primary cohort analysis, and in stratified comparisons of ICU vs non-ICU patient sera 3 weeks or more after symptom onset.

This is the first study to simultaneously measure SARS-CoV-2 antibody levels to the RBD and NP using an automated immunoassay in longitudinal serum samples. Previous reports have used research grade ELISAs that require manual dilutions to obtain a semi-quantitative titer or signal to cut-off or calibrator ratio,^5,12,14^ except for one automated magnetic chemiluminescence enzyme immunoassay.^6^ Time to seroconversion estimates are dependent on the sensitivity of the analytical method and the viral antigen used for measurement, as well as the length of follow-up for the cohort. Similarly, differences in the analytical dynamic range between SARS-CoV-2 antibody methods and follow-up times can affect the validity of peak measurements, confounding direct comparisons between critical and non-critical cases. These differences among SARS-CoV-2 antibody methods and study designs likely account for discrepant findings reported to date and should be considered when evaluating future studies. Some new SARS-CoV-2 antibody assays run on automated analyzers are positioned as qualitative and may be less informative to evaluate antibody kinetics and correlation with disease severity or neutralizing ability.

Most COVID-19 serological studies have focused on critical hospitalized cases. Quantitative SARS-CoV-2 IgG data from recovered patients with mild disease are sparse and lack correlative data to severe disease from the same time intervals post symptom onset. Here we report that among 63 SARS-CoV-2 RT-PCR positive patients with non-critical disease tested >3 weeks after the onset of symptoms, 54 (85.7%) had detectable levels of SARS-CoV-2 IgG. However, IgM and IgG were significantly lower than the corresponding data for ICU patients in the same timeframe since symptom onset. Studies of people infected with SARS-CoV and MERS suggest that the antibody response wanes over time but is detectable more than one year after hospitalization.^15-17^ It is too soon to determine if the SARS-CoV-2 IgG response will persist, especially in mild cases with a limited antibody response.

The extent to which a robust antibody response to SARS-CoV-2 results in virus neutralization or contributes to the pathology in severe COVID-19 disease is still unknown.^18^ Antibody concentrations generally correlate with host protection and viral neutralization, but in rare cases antibodies can promote disease progression, resulting in a phenomenon known as antibody-dependent enhancement (ADE).^19^ For SARS-CoV, ADE was shown to promote virus uptake into macrophages resulting in elevated production of inflammatory cytokines and acute lung injury.^20,21^ Higher levels of SARS-CoV-2 antibodies in severe cases in our study could suggest a similar mechanism for COVID-19.^19^ In a small study of serum samples taken from 16 patients 14 days or longer after symptom onset, anti-SARS-Cov-2-NP and anti-SARS-CoV-2-RBD IgG levels correlated with virus neutralization titer.^3^ A recent investigation of patients recovered from mild to moderate COVID-19 demonstrated that anti-RBD IgG levels correlated well with CD4+ T cell responses to the spike protein, raising optimism for some durability of immunity and the potential for vaccine efficacy.^22^ More research is key to characterize SARS-CoV-2 antibody avidity and neutralization ability, and to determine both beneficial and potentially pathogenic outcomes associated with the magnitude of the antibody response.

Our data have important implications for the use of conventional IgG serology as a tool to address the COVID-19 pandemic. Population-based IgG seroprevalence may underestimate the occurrence of mild infections, with the degree of underestimation dependent on the sensitivity of the screening method. Rapid tests with reported sensitivity of 90% in hospitalized patients may have substantially lower sensitivity in sera from mild or asymptomatic infections. High sensitivity assays like the one we employed, with robust high to low dynamic range, can provide a more complete picture of cumulative incidence. Underestimation in population surveys that use rapid screening tests could be gauged by testing a representative sample of rapid test-negative specimens with a high sensitivity assay. A comprehensive understanding of the correlates of protective immunity, both cellular and humoral, is even more crucial. Such an understanding will provide a necessary foundation for the therapeutic use of convalescent plasma, predictive epidemiological modeling and projections of vaccine effectiveness.

## Data Availability

The data can be made available via request to the corresponding author.

